# SARS-CoV-2 serosurvey across multiple waves of the COVID-19 pandemic in New York City between 2020-2023

**DOI:** 10.1101/2023.12.18.23300131

**Authors:** Juan Manuel Carreño, Abram L. Wagner, Brian Monahan, Daniel Floda, Ana S Gonzalez-Reiche, Johnstone Tcheou, Ariel Raskin, Dominika Bielak, Gagandeep Singh, Sara Morris, Miriam Fried, Temima Yellin, Leeba Sullivan, PARIS study group, Emilia Mia Sordillo, Aubree Gordon, Harm van Bakel, Viviana Simon, Florian Krammer

## Abstract

Sero-monitoring provides context to the epidemiology of severe acute respiratory syndrome coronavirus 2 (SARS-CoV-2) infections and changes in population immunity following vaccine introduction. Here, we describe results of a cross-sectional hospital-based study of anti-spike seroprevalence in New York City (NYC) from February 2020 to July 2022, and a follow-up period from August 2023 to October 2023. Samples from 55,092 individuals, spanning five epidemiological waves were analyzed. Prevalence ratios (PR) were obtained using Poisson regression. Anti-spike antibody levels increased gradually over the first two waves, with a sharp increase during the 3^rd^ wave coinciding with SARS-CoV-2 vaccination in NYC resulting in seroprevalence levels >90% by July 2022. Our data provide insights into the dynamic changes in immunity occurring in a large and diverse metropolitan community faced with a new viral pathogen and reflects the patterns of antibody responses as the pandemic transitions into an endemic stage.

## Introduction

Coronavirus disease 2019 (COVID-19) has severely impacted human health worldwide. Within the United States, New York City emerged as an early epicenter of infection and cases, with the first reported case on February 29, 2020^1^. However, public reporting of cases over time was inconsistent due to limited testing capacity early in the pandemic^1^. Moreover, testing was differentially available to groups depending on their socioeconomic status^2^, impacting understanding of the course of the epidemic in different populations.

Severe acute respiratory syndrome coronavirus 2 (SARS-CoV-2), the causative agent of COVID-19, has rapidly evolved to increase its transmission potential and to evade immunity derived from prior infection and vaccination, contributing to several waves of infections^3^. The emergence of viral variants capable of overcoming pre-established immune responses is mainly attributed to mutations in the spike surface glycoprotein. During the first wave of infections, the wild type (WT) virus acquired the D614G mutation that increased virus infectivity and transmissibility^4, 5^. These strains prevailed for several months until the second wave of infections caused by the Alpha, Beta and Gamma variants occurred in different parts of the world. A 3^rd^ wave of infections was caused by the Delta variant which showed higher transmissibility and enhanced severe disease. A significant reduction of viral neutralization by sera from convalescent or vaccinated individuals was detected^6, 7^. In December 2021, the emergence of the Omicron BA.1 strain, which carried over 30 spike protein mutations, resulted in a 4^th^ wave of infections. Since then, distinct Omicron sub-lineages have circulated worldwide, including BA.2, BA.5, BQ.1, BQ.1.1, BF.7, and more recently XBB.1, XBB.1.5, BA.2.86, EG.5.1, JN.1, and derived variants.

Throughout the pandemic, monitoring the level of immunity in the population has been crucial for public health. Sero-monitoring has guided safety measures and vaccination policies worldwide to reduce viral infection, transmission, and severe disease^8^. Laboratories can measure binding antibodies to the spike (S) protein, which indicates past infection or vaccination, or antibodies to the nucleoprotein (NP), which indicates a past infection. The level of anti-SARS-CoV-2 antibodies and T cell responses correlate with an individual’s capacity to combat the virus and with protection against infection and disease^9, 10^. Importantly, binding antibody levels correlate with the number of prior exposures, and with the level of neutralizing antibodies^11, 12^. Hence, sero-surveillance for SARS-CoV-2 provides an estimate of the level of immunity in the population and reflects the patterns of the pandemic. Moreover, existing case reporting systems for COVID-19 are limited by changing testing availability and behaviors over time.

Here, we report the results of a prospective, hospital-based cross-sectional study of anti-S and anti-NP sero-monitoring from the beginning of February 2020 through July 2022, and follow-up measurements in August-October 2023. We describe the longitudinal seroprevalence in two different groups: a) an ‘urgent care’ group that was enriched for acute cases of COVID-19 early during the pandemic, and b) a ‘routine care’ group, resembling the general population, as previously described^8^. Stratification by zip code allowed us to study the geographical distribution of the seroprevalence and antibody titers in residents of NYC throughout the pandemic.

## Methods

### Study participants and human samples

Sample collection and testing started on February 9^th^, 2020 and ended on July 18^th^, 2022. Follow up testing started on August 28^st^ 2023 and ended on October 2^nd^ 2023. Collection was performed on a weekly basis (n=608/week on average) and in a blinded manner. Plasma was collected from ethylenediaminetetraacetic acid (EDTA)-treated blood specimens that remained from standard-of-care testing at the Mount Sinai Hospital (MSH) Blood Bank. Samples were sorted to collection week, visit location and practice including: OB/GYN, labor and deliveries; oncology; surgery; cardiology; emergency department; and other related hospital admissions. Up to 152 specimens at each location in a defined collection week were randomly selected.

During sample aliquoting, specimens were captured into a coded database with randomly assigned identifiers and verified patient categories. Additional specimens obtained from the same patient within the same week were not considered for analysis. Samples collected 7 days apart or more from the first specimen collected were considered as independent samples of the population. Typically, 67-314 specimens were selected weekly from the ‘routine care’ group (OB/GYN, labor and deliveries, oncology, surgery, and cardiology) and 67 to 314 samples from the ‘urgent care’ group consisting of specimens from patient visits to the emergency department and urgent care. Some individuals had a PCR test to diagnose SARS-CoV-2 infection. The analyses of samples collected during the week ending on February 9^th^ 2020 to July 5^th^ 2020 were reported previously^8^. We incorporate all samples with additional demographics data in the current analysis in order to describe seroprevalence and antibody titers since the beginning of the pandemic. We stratify our analyses by ‘urgent care’ vs ‘routine care’ groups to capture the fact that access to non-urgent medical care was limited at the beginning of the pandemic^13^.

Specimen metadata included information on the individuals’ age, sex, race, insurance status, and whether they had received a COVID-19 vaccination prior to specimen collection. Due to the low number of children in our study, we limit our main analysis to adults, and divided them into 18-44 years, 45-64 years, and 65+ years age groups. Information on race was based on self-reported categories (e.g., Asian and Pacific Islander, African American, White, Other, and unknown). Information on ethnicity (e.g., Hispanic or non-Hispanic) was not consistently available and, thus, not included in our analysis. Medicaid and Medicare were collapsed into a public insurance category. All data used in this study was anonymized and recoded through the use of an honest broker following local regulations.

### Epidemiological waves

We categorized samples by time into 5 epidemiological waves, that correspond to successive peaks in COVID-19 incidence in NYC: Wave 1 (February 9^th^ to August 30^th^, 2020; ancestral SARS-CoV-2 and D614G mutant circulating), Wave 2 (August 31^st^, 2020, to June 20^th^, 2021; Iota (B.1.529) and Alpha (B.1.1.7) circulating), Wave 3 (June 21^st^ to October 31^st^, 2021; Delta (B.1.617.2) circulating), Wave 4 (November 1^st^, 2021, to March 6^th^, 2022; Delta (B.1.617.2) and Omicron BA.1 (B.1.1.529.1) circulating), and Wave 5 (March 7^th^ to July 18^th^, 2022; Omicron BA.2 (B.1.1.529.2) and Omicron BA.5 (B.1.1.529.5) circulating). Epidemiological waves and circulating variants are depicted in Figures 1A and 1B respectively. The follow up period (August 28^st^ 2023 - October 2^nd^ 2023), was not associated to a specific wave timeframe, and was included to assess the maintenance of seroprevalence and durability of spike binding antibody titers.

**Figure 1.**
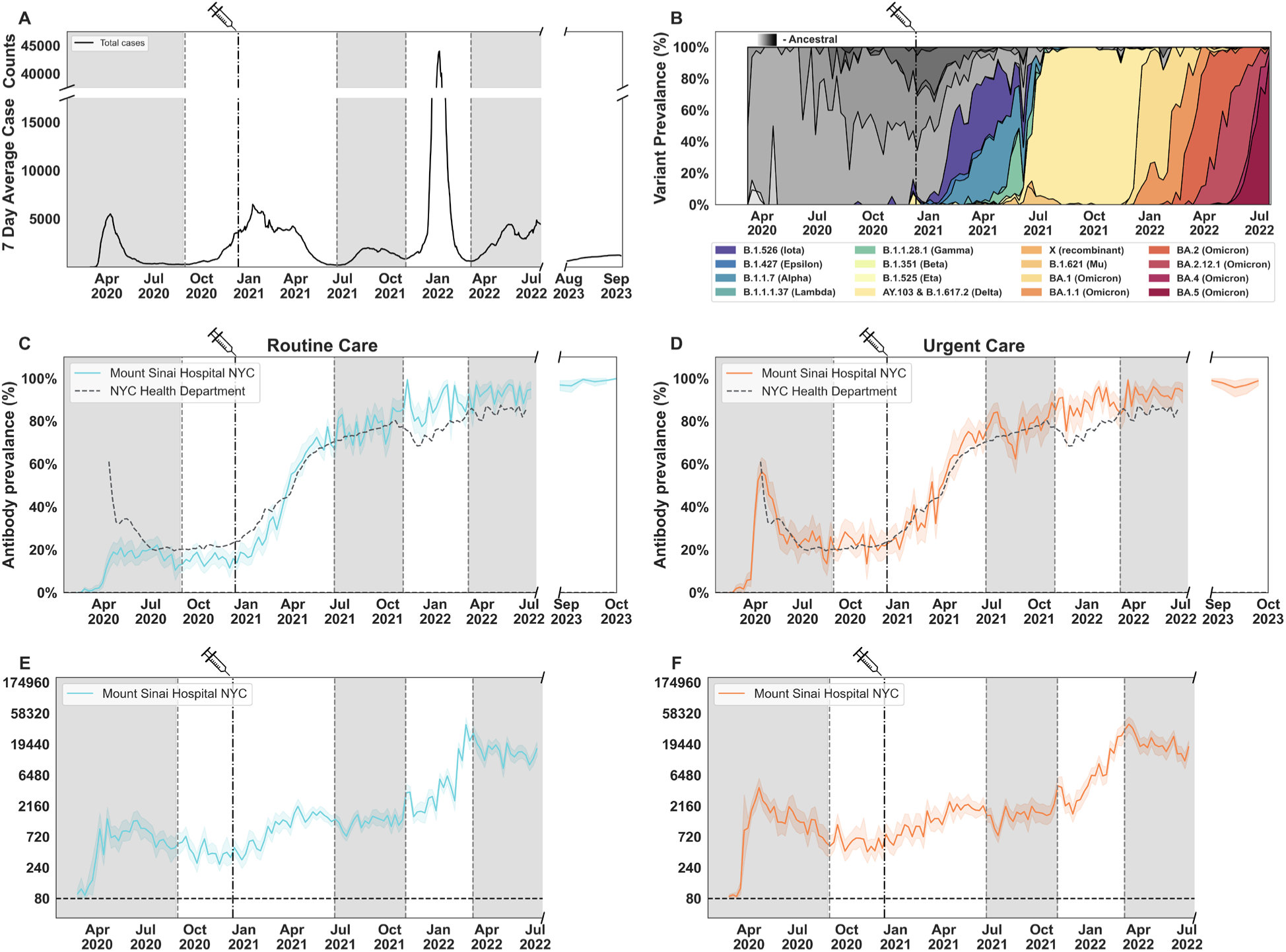
SARS-CoV-2 spike binding antibody prevalence and titers in the Routine care and Urgent care groups at a Mount Sinai Hospital in New York City (NYC). Average case counts (7-day) of SARS-CoV-2 infections in New York City reported by the NYC Department of Health and Mental Hygiene (NYC DOHMH) (**A**). Main circulating SARS-CoV-2 lineages based on data from the Pathogen Surveillance Program at Mount Sinai (**B**). Antibody prevalence (**C, D**) and titers (**E, F**) in the Routine Care (left column, blue lines) and Urgent Care (right column, orange lines) groups, measured between February 9^th^ 2020 to July 18^th^ 2022, and seroprevalence measured between August 21^st^ 2023 to October 2^nd^ 2023. Charts are fitted with a bootstrapped confidence interval based on random resampling of results to compute an estimate of the 95% confidence interval of the mean. In (**B**), lineages before the emergence of variants of concern (VOCs) are shown in greyscale, while VOCs are shown in color. NYC DOHMH antibody prevalence data is shown in **C-D** for reference, denoted with a dashed line. The date on which the first FDA-authorized SARS-CoV-2 vaccine became available in NYC is indicated by the vertical doted line and syringe. Alternating shaded areas in **C-F** denote the five successive epidemiological waves of infection in NYC.

### Geographical maps

The neighborhood tabulation area (NTA) map created by the Department of City Planning and sourced from NYC OpenData was used as the base geometry for our data analyses. For each NTA, the average of the common logarithm of spike titer values was calculated and plotted as a shaded map using the ggplot2 and sf packages in R. For plots showing progression over time, slices were taken from each data range and plotted as separate facets.

### Statistical analysis

We describe the distribution of results graphically and through proportions, stratified by ‘urgent care’ vs ‘routine care’ groups. Visualizations of antibody prevalence and titers were generated using Matplotlib (3.7.2), Seaborn (0.11.2), Pandas (1.5.3), Numpy (1.26.0), and Scipy (1.11.3) packages within python 3.11.5. Geographical charts, and live data explorer were generated using R version 4.3.2, with the packages sf (1.0.14), DT (0.31), zoo (1.8.12), Hmisc (5.1.1), shiny (1.8.0), scico (1.5.0), ggpubr (0.6.0), bsplus (0.1.4), forcats (1.0.0), ggplot2 (3.4.4), ggbreak (0.1.2), patchwork (1.1.3), lubridate (1.9.3), hrbrthemes (0.8.0), data.table (1.14.10), shinyWidgets (0.8.0), RColorBrewer (1.1.3), shinydashboard (0.7.2), and shinycssloaders (1.0.0).

Based on *a priori* considerations, we include age, sex, race, and insurance status in our multivariable models: age representing differential humoral response to infection and increased risk of severe disease^14, 15^ and therefore a marker of potentially enhanced social distancing behaviors^16^, sex linked to health seeking behaviors^17^ as well as gynecological appointments, and race and insurance status measures of poverty, health seeking behaviors, and exposure to SARS-CoV-2 through a variety of mechanisms^18^.

Differences in seropositivity by demographic group over time were assessed through multivariable, Poisson regression models that included robust variance estimates^19^. These models output prevalence ratios (PR) and 95% confidence intervals (CI), which were interpreted relative to a reference group. These models estimated the direct effect of age, sex, race, and insurance status on seropositivity. These models are initially stratified by epidemiological wave. To estimate changes in seropositivity by demographic group across wave, we also specified a larger model, where an interaction term was placed between wave and every demographic variable. The p-value from this interaction term indicates if there has been a significant shift in differences across wave, for a given demographic variable.

We conduct several sensitivity analyses. In a separate analysis, we also include vaccination, but limit the timing to data collected after January 11, 2021, when the vaccine started becoming available to certain age groups in the general population^20^. And in a third set of models, we exclude individuals 65+ based on their ability to access Medicare. The regression models were implanted in SAS version 9.4 (SAS Institute, Cary, NC, USA). We used an alpha level of 0.05 to assess significance.

### Ethical approval

The study protocol HS 20-00308 was reviewed by the Mount Sinai Health System Institutional Review Board, Icahn School of Medicine at Mount Sinai, and it was exempt from human research as defined by regulations of the Department of Health and Human Services (45 CFR 46. 104).

### Data availability

All data are available under ImmPort accession #xxx.

## Results

Overall, there are 55,092 individuals sampled between February 9, 2020 and July 18, 2022: 21,075 from urgent care group and 34,017 from the routine care group. Of these 882 were children <18, and 54,210 were adults. The demographic distribution among adults is shown in **Table 1**. In the routine care groups, a plurality of individuals were in the 18-44 year age group (16,499, 49%), a majority female (23,155, 69%), a plurality self-identified as White (14,971, 45%), and about half had private insurance (16,386, 49%). The distribution of these variables for children is shown in **Supplementary Table 1**. COVID-19 vaccination began in January 2021 in the general population and varied across a number of groups, including race. Uptake was faster in Asian and Pacific Islanders and White New Yorkers compared to African Americans (e.g., at wave 3, uptake in these three groups was 63%, 55%, and 50%, respectively) (**Supplementary Table 5**).

We measured spike antibody prevalence and titers over five epidemiological waves based on average case counts (7-day) of SARS-CoV-2 infections in New York City, reported by the NYC Department of Health and Mental Hygiene (NYC DOHMH) (**Figure 1A**). Viral variant prevalence through this period continuously changed, from the initial circulating of ancestral variants to the appearance of variants of the Omicron lineage (**Figure 1B**). Overall, among adults in the routine care group, spike protein seropositivity increased from an average of 13% in wave 1, to 36%, 76%, 88%, and 93% in the following four waves (**Figure 1, C-D; Supplementary Table 2**), consistent with seroprevalence estimates reported by the NYC Department of Health and Mental Hygiene (DOHMH)^21^. A sharp increase in antibody prevalence was detected during waves 2 and 3, starting on February 2021. This increase was corresponded temporally with expanded availability of SARS-CoV-2 vaccinations, the emergence of the Iota, Alpha, and Delta variants, and changes in public health measures. Importantly, antibody prevalence was maintained during the follow up period from August 28^st^ 2023 to October 2^nd^ 2023 (**Figure 1, C-D**). Vaccinated individuals showed significantly higher antibody prevalence than non-vaccinated as early as January 2021, with the largest differences just prior to and during wave 3 (**Figure 2, G-H**) owing to incomplete vaccination in the earlier periods and unvaccinated individuals experiencing high infection rates during wave 3. By wave 5, spike protein seropositivity was uniformly high (≥90% in all categories). Similar trends were observed in populations stratified by gender and age, with no significant differences among intra-group strata (**Figure 2, A-D**). Differences by race were observed, for example, 14% of African Americans and 18% of those within the Other racial category were seropositive in wave 1, versus 7% of Asians and Pacific Islanders (**Figure 2, E-F**).

**Figure 2.**
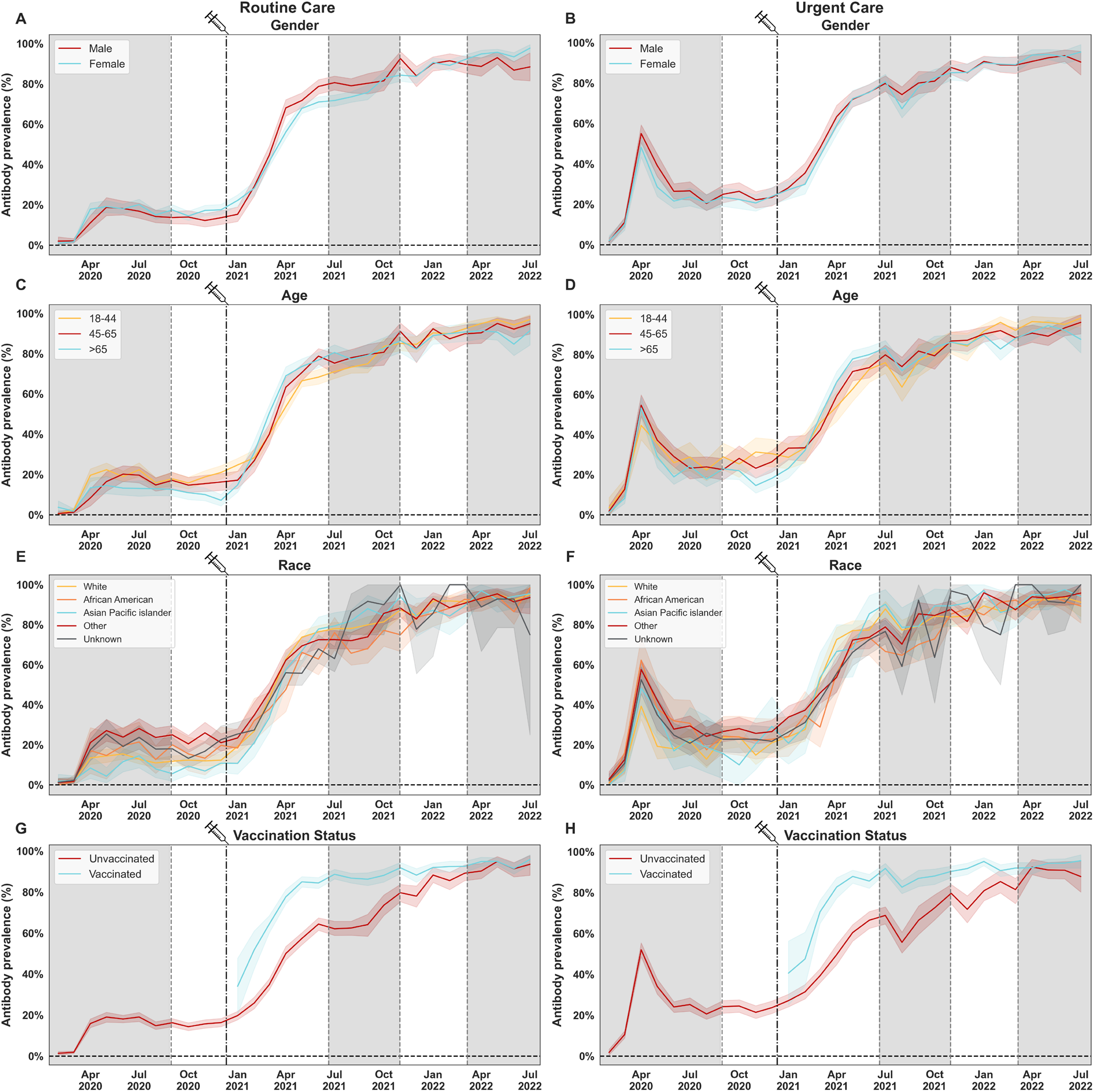
SARS-CoV-2 spike antibody prevalence stratified by demographic groups and vaccination status. Antibody prevalence in the Routine Care (left column), and Urgent Care (right column) groups, measured between February 9^th^ 2020 to July 18^th^ 2022, and stratified by gender (**A, B**), age (**C, D**), race (**E, F**), and vaccination status at the time of sample collection (**G, H**) is shown. Charts are fitted with a bootstrapped confidence interval based on random resampling of results to compute an estimate of the 95% confidence interval of the mean. **A, B.** Gender stratification: males, females. **C, D**. Categorical age levels: 18-44, 45-65, >65. Individuals <17 are not shown in this analysis. **E, F**. Race stratification: White, African American, Asian and Pacific islander, Other, unknown. **G,H.** The date on which the first FDA-authorized SARS-CoV-2 vaccine became available in NYC is indicated by the vertical doted line and syringe. Vaccination status was assessed at time of sample collection and does not reflect vaccination rates in NYC or within our patient population.

The multivariable Poisson regression model of spike protein seropositivity reveals several trends within demographic group and across time (**Table 2**). At wave 1, seropositivity was higher in younger age groups compared to older (1.66 times higher in those 18-44 vs 65+, 95% CI: 1.41, 1.96). By wave 5, this had largely attenuated (interaction *p-*value <0.0001). Larger disparities by race were also observed at the beginning of the pandemic. Compared to White New Yorkers, Black (prevalence ratios (PR): 1.24, 95% CI: 0.94, 1.63) and Other New Yorkers (PR: 1.64, 95% CI: 1.44, 1.88) had higher seropositivity, and Asians and Pacific Islanders had relatively low seropositivity (PR: 0.67, 95% CI: 0.51, 0.87). During waves 3 and 4, some of these associations reversed, with lower seropositivity in Black and Other groups compared to White New Yorkers. Around a similar or earlier time (waves 2 and 3), vaccination coverage was relatively low among Black compared to White New Yorkers (Supplementary Table 5). An analysis of spike protein seropositivity across racial ethnic groups stratified by vaccination status reveals significant differences among the unvaccinated (Supplementary Table 6), with seropositivity relatively high in waves 1 through 2 and then relatively low in waves 3 and 4 in Black New Yorkers. However, among the vaccinated, seropositivity remained equivalent across racial groups. Indeed, in the models with vaccination status that cover periods once the vaccine had been rolled out (**Supplementary Table 3**), this factor emerges as a significant contributor to seropositivity, with a larger impact in wave 2, where seropositivity was 1.95 times higher among those vaccinated compared to unvaccinated (95% CI: 1.87, 2.03), while it was only 1.04 times higher in wave 5 (95% CI: 1.02, 1.07) (interaction *p-*value <0.0001).

Anti-spike antibody titers in seropositive individuals were induced at moderate to high levels during the first waves of infections, with modest antibody decay over time (**Figure 1, E-F; Supplementary Figure 1**). Amid the second wave, titers increased gradually reaching the highest levels during wave 5. No significant differences were detected when titers were stratified by gender, age or race (**Figure 3, A-F**). As expected, the introduction of vaccination during wave 2 resulted in increased antibody prevalence and titers at the time of sample collection, while individuals without a record of vaccination displayed significantly lower antibody prevalence through waves 2, 3 and 4 (**Figures 2 and 3, G-H**). These differences narrowed over time, likely due to multiple infections in unvaccinated individuals. Moreover, differences in antibody prevalence and titers in vaccinated vs non-vaccinated individuals were preserved when stratification was based on sex, age, and race (**Supplementary Figures 2-7**).

**Figure 3.**
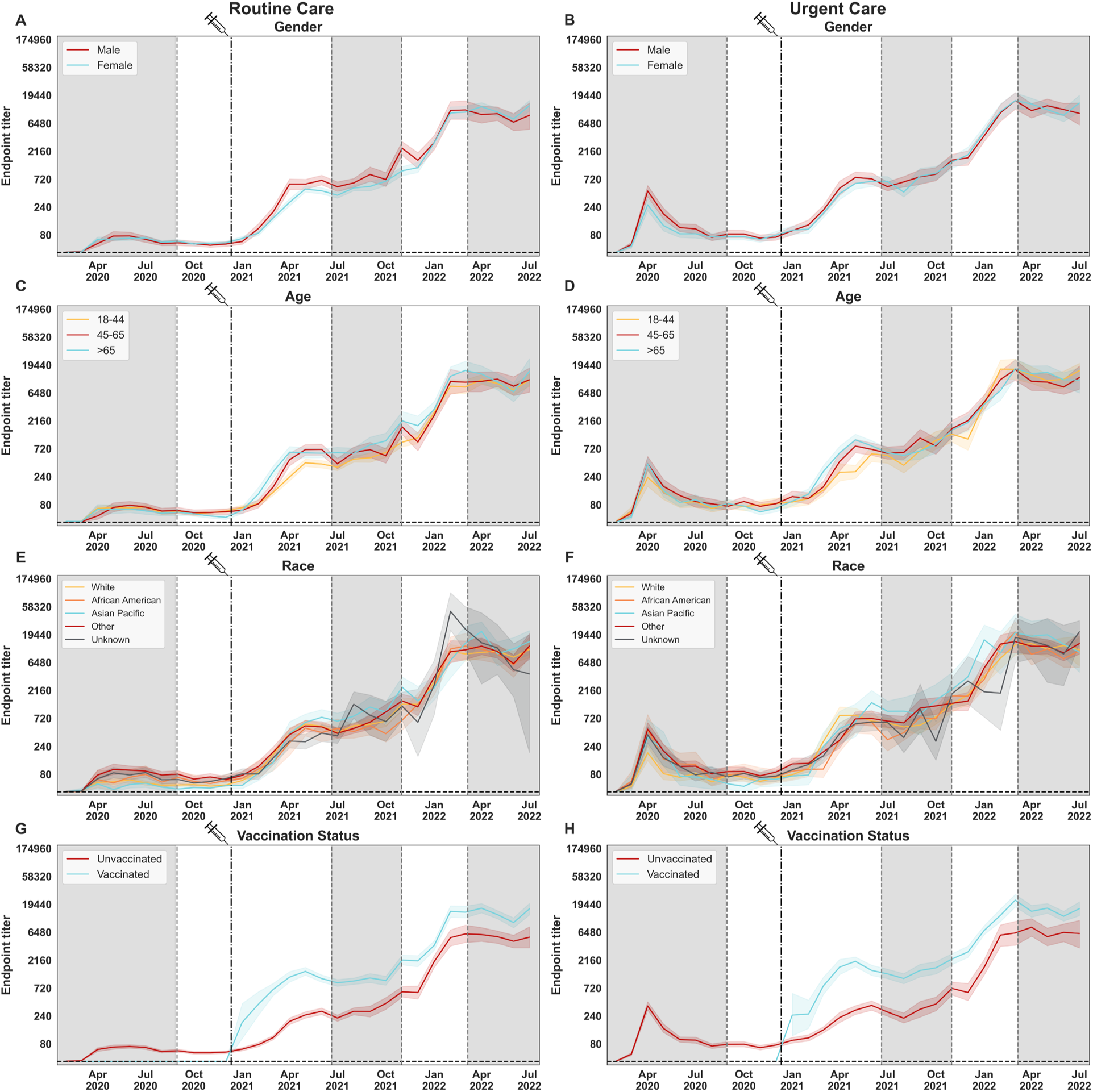
SARS-CoV-2 spike antibody titers by demographic groups and vaccination status. SARS-CoV-2 spike antibody prevalence in the Routine Care (left column), and Urgent care (right column) groups measured between February 9^th^ 2020 to July 18^th^ 2022 stratified by gender (**A, B**), age (**C, D**), race (**E, F**), and vaccination status at the time of sample collection (**G, H**). Charts are fitted with a bootstrapped confidence interval based on random resampling of results to compute an estimate of the 95% confidence interval of the mean. **A, B.** Gender stratification: males, females. **C, D**. Categorical age levels: 18-44, 45-65, >65. Individuals <17 not shown in this analysis. **E, F**. Race stratification: White, African American, Asian and Pacific islander, Other, unknown. **G,H.** The date on which the first FDA-authorized SARS-CoV-2 vaccine became available in NYC is indicated by the vertical doted line and syringe. Vaccination status was assessed at time of sample collection and does not reflect vaccination rates in NYC or within our patient population.

For select weeks (July 6^th^-July 14^th^ 2020, February 15^th^-March 1^st^ 2021, June 1^st^ – June 14^th^ 2021, August 16^th^ – August 30^th^ 2021, May 23^rd^ – May 30^th^ 2022, August 28^th^ – Oct 2^nd^ 2023) we also measured seropositivity and titers for nucleoprotein (NP). Although initially in 2020 NP seropositivity was similar to the spike seropositivity, this changed in 2021 as NP seropositivity fell behind spike positivity by a relatively large margin (**Figure 4**). This was influenced not only by vaccination (which only induces anti-spike antibodies) but also by waning of NP antibodies over time and the low induction of NP antibodies during breakthrough infections. Our estimates during mid 2021 (June 1^st^ – June 14^st^, 57% (95% CI = 53.1–60.9) to late 2021 (August 16^th^ – August 30^th^, 54% (95% CI = 50.5–57.5)) are higher than the reported nationwide NP seroprevalence (December 2021 33.5% (95% CI = 33.1–34.0))^22^. These differences may be due to discrepancies in the time points of antibody measurements. Antibodies induced or boosted during the Delta wave may have waned over the subsequent months, due to the relatively fast decay rates of NP antibodies induced by infection^11^. Alternatively, differences in the sensitivity of the methods used or higher COVID-19 incidence in NYC compared to nationwide levels^23^ could have played a role.

**Figure 4.**
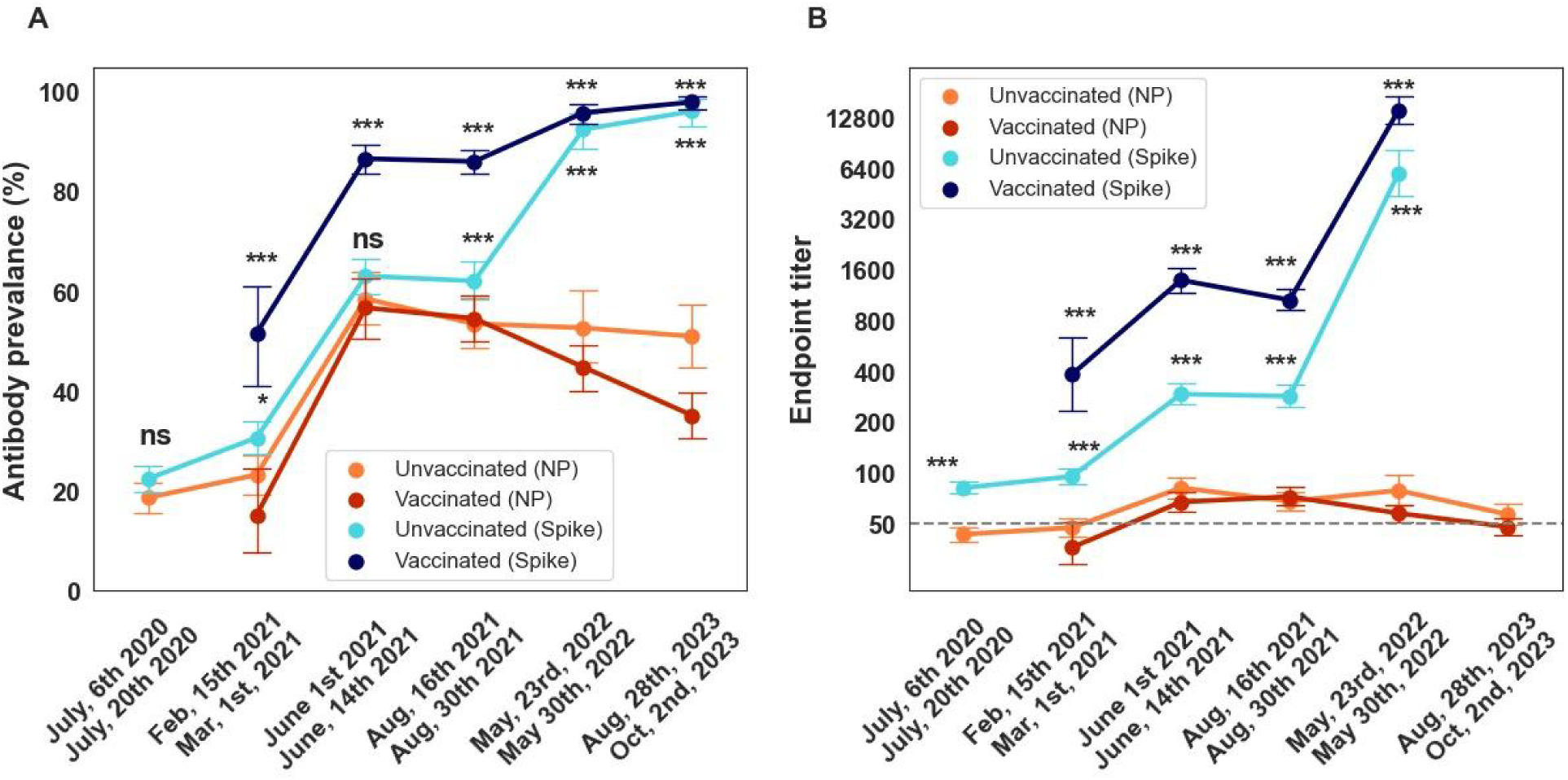
SARS-CoV-2 nucleoprotein (NP) and spike (S) antibody prevalence and titers over six sampling periods (2-week duration) through the course of the pandemic. NP and S antibody prevalence (**A**) and titers (**B**) in patients attending a Mount Sinai Hospital in New York City are shown. Data stratified by vaccination status, assessed at the time of sample collection. Sampling time points: July 6^th^ - July 20^th^, 2020; February 15^th^ - March 1^st^, 2021; June 1^st^ - June 14^th^, 2021; August 16^th^ – August 30^th^, 2021; May 23^rd^ - May 30^th^, 2022; Aug 28^th^ – Oct 2^nd^, 2023. Antibody prevalence (%) plus 95% confidence intervals are shown in **A**. Geometric mean of endpoint titers plus 95% confidence intervals are shown in **B**. Limit of detection (LoD) is indicated by the horizontal dotted line. Statistically significant differences between NP and S values are indicated as follows: ns, not significant; * <0.05, ** <0.01, *** <0.001 (Chi squared test for antibody prevalence, and T test for Endpoint titer)"

To analyze the geographical distribution of antibody prevalence and titers through the five boroughs of NYC (Manhattan, Brooklyn, Queens, The Bronx, and Staten Island), we used neighborhood tabulation areas (NTAs) to build maps of the city divided into 216 NTAs (**Figure 5**). We detected higher antibody prevalence in areas located outside Manhattan during all waves of infections, that were more evident during waves 1-3, when antibody prevalence was lower (**Figure 5, A-C**). Importantly, seroprevalence reached high levels (above 80%) almost homogenously throughout the five boroughs by wave 5 (**Figure 5, E**). Likewise, higher titers were detected in boroughs other than Manhattan (**Figure 6**), with areas reaching the highest titers located in Queens (**Figure 6, E**).

**Figure 5.**
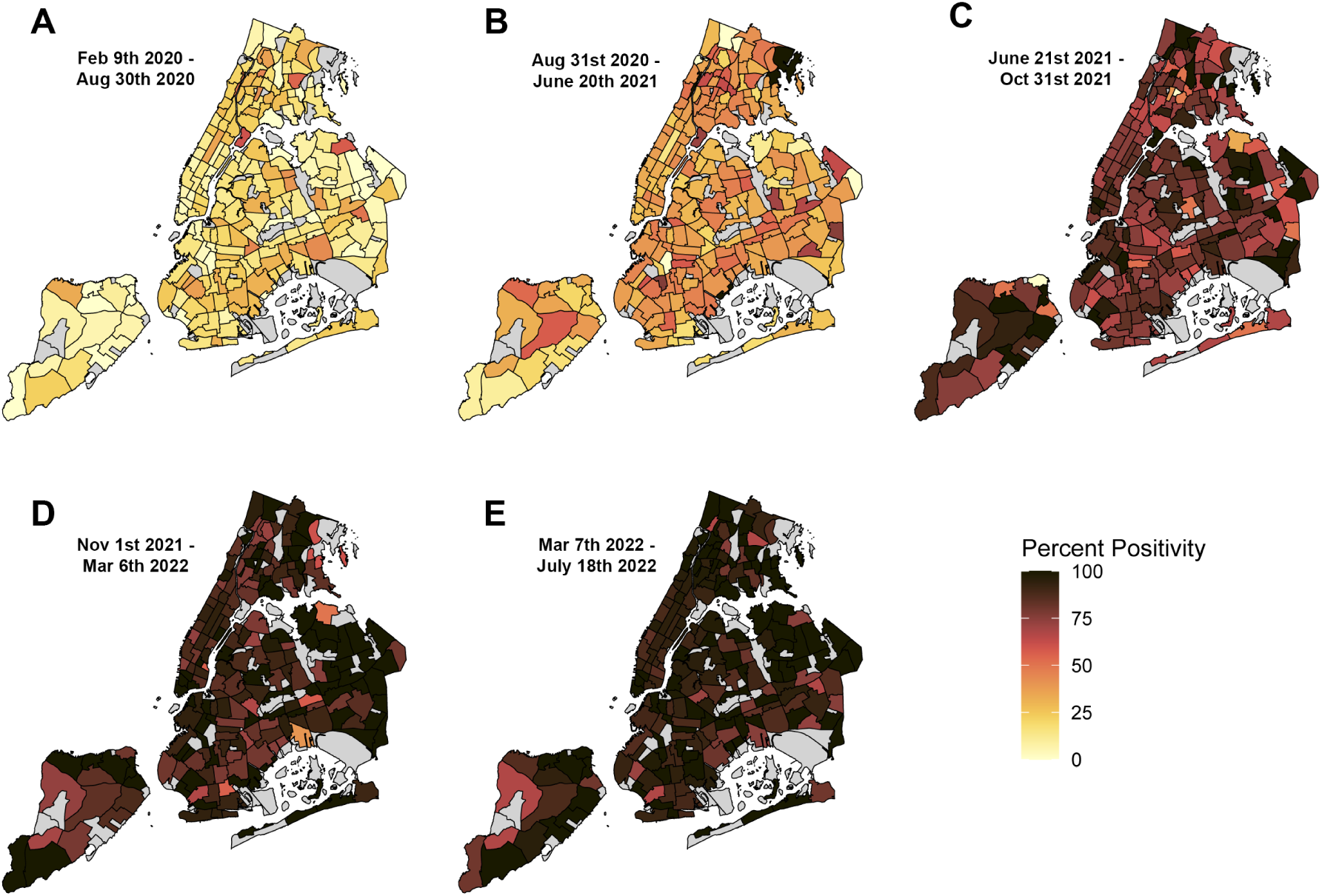
Geographical distribution of SARS-CoV-2 spike antibody prevalence during five epidemiological waves of COVID-19 in residents of New York City (NYC). SARS-CoV-2 spike IgG prevalence measured in patients from the five boroughs of NYC (Manhattan, Brooklyn, Queens, The Bronx, and Staten Island) attending to a Mount Sinai Hospital in NYC are shown. 5 epidemiological waves corresponding to successive peaks of COVID-19 incidence in NYC are shown. **A**. Wave 1 (February 9^th^ to August 30^th^, 2020). **B**. Wave 2 (August 31^st^, 2020, to June 20^th^, 2021). **C**. Wave 3 (June 21^st^ to October 31^st^, 2021). **D**. Wave 4 (November 1^st^, 2021, to March 6^th^, 2022). **E**. Wave 5 (March 7^th^ to July 18^th^, 2022). Antibody prevalence is expressed as percent positivity within neighborhood tabulation areas. Areas with less than 10 specimens are shaded grey. Color gradient depicts percent positivity (0 to 100%).

**Figure 6.**
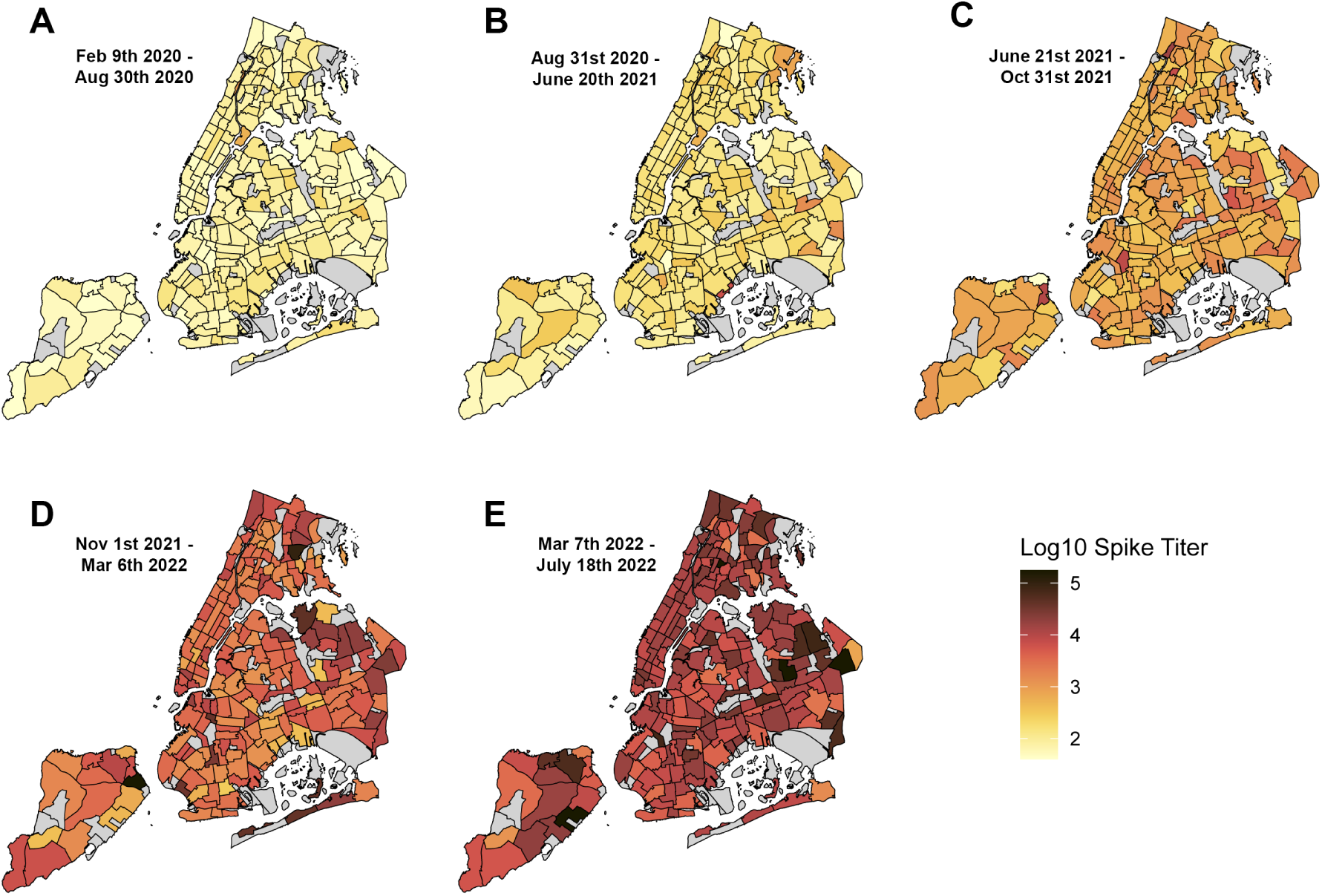
Geographical distribution of SARS-CoV-2 spike antibody titers during five epidemiological waves of COVID-19 in residents of New York City (NYC). SARS-CoV-2 spike IgG titer measured in patients from the five boroughs of NYC (Manhattan, Brooklyn, Queens, The Bronx, and Staten Island) attending to a Mount Sinai Hospital in NYC are shown. 5 epidemiological waves corresponding to successive peaks of COVID-19 incidence are shown. **A**. Wave 1 (February 9^th^ to August 30^th^, 2020). **B**. Wave 2 (August 31^st^, 2020, to June 20^th^, 2021). **C**. Wave 3 (June 21^st^ to October 31^st^, 2021). **D**. Wave 4 (November 1^st^, 2021, to March 6^th^, 2022). **E**. Wave 5 (March 7^th^ to July 18^th^, 2022). Antibody titer is expressed as log10 geometric mean titer. Areas with less than 10 specimens are shaded grey. Color gradient depicts range of antibody titers.

## Discussion

Serosurveillance systems provide means to describe population patterns in infection and vaccination over time. Overall, in a hospital-based study in New York City, we previously described a sharp increase in anti-spike antibody prevalence early during 2020 due to SARS-CoV-2 infections^8^. By May 2021, seroprevalence reached levels above 70% due to both infections, and vaccination programs. Here we report on the continued sero-monitoring efforts to capture antibody prevalence and titers over the course of the pandemic. We show that antibody prevalence gradually increased over time, reaching levels above 90% by January 2022. Seroprevalence and titers were maintained over time, with similar trends observed during a follow up period in August-October, 2023, likely due to constant antigenic exposures by vaccine boosters and breakthrough infections. Moreover, we analyzed the geographical distribution of antibody prevalence and titers through the five boroughs of NYC. Finally, we analyzed NP titers – indicative of immunity derived by infection – at discreet timepoints during the pandemic.

Our seroprevalence estimates for the most part overlap with estimates from the NYC DOHMH. However, there are differences during the early phase of the pandemic and later during waves 4 and 5. Specifically, the NYC DOHMH reports a large proportion (>50%) of seropositive individuals in wave 1 who were tested for SARS-CoV_2 by PCR. Individuals seeking testing were probably more likely to be infected and potentially already antibody positive given the long initial incubation time^1^. This is similar to what we observed in the urgent care group that was biased towards SARS-CoV-2 positives in the first wave. In more recent waves, seropositivity is lower according to the NYC DOHMH than in our serosurveillance study. This could reflect differences in the assays used or differences in the study populations.

The COVID-19 pandemic intensified racial and economic disparities in health^24^. Our data collected in the early waves, indicate that seropositivity was relatively high in adults self-identifying as Black and Others, pointing to higher infection rates in essential workers lacking the possibility to socially distance or due socioeconomic constraints leading to more crowded living conditions^25, 26^. Our serosurveillance system revealed other sociodemographic trends by age. These reflect epidemiological risk patterns and vaccination policies. For example, early in the pandemic, seropositivity was greater in younger adults, in accordance with risk messaging directed towards older adults. As vaccination became available and was prioritized for older ages^27^, seropositivity became relatively higher in older age groups.

The spike antibody response induced towards the emerging Omicron sub-lineages is mostly cross-reactive with ancestral strains^28^, hence the seroprevalence estimates presented here are likely to be preserved towards the spike of Omicron sub-variants. Although the neutralization capacity of sera from individuals exposed to ancestral SARS-CoV-2 antigens through infection or vaccination is severely reduced against Omicron strains^29^, non-neutralizing antibodies may protect through alternative mechanisms mediated by Fc-dependent antibody effector functions^30^ and the presence of antibodies likely also indicates the presence of T-cell responses which largely cross-react between different variants of concern ^31^. Dissecting the contribution of non-neutralizing cross-reactive antibodies upon exposure to emerging variants of the Omicron lineage will be important to have a broader picture of an individual’s immunity.

Our analysis has some limitations. Our catchment area was tied to individuals visiting the Mount Sinai Health System, and we may have limited scope in generalizing outside the NYC metropolitan area. Residual, de-identified samples are collected, which impedes following individuals longitudinally. Representation of samples by different zones was limited, which limited the geographical distribution analysis.

## Conclusions

New York City was an early epicenter of the COVID-19 pandemic. Leveraging a hospital-based serosurveillance system, we detect several patterns in line with the epidemiology of SARS-CoV-2 and the roll-out of the COVID-19 vaccines. Notably, by early 2022, virtually the entire population had some evidence of COVID-19 seropositivity. Importantly our data clearly show increasing overall seropositivity in the general population as SARS-CoV-2 vaccines became available. The overall sustained high level of implied immunity in the total population also is noteworthy. Finally, we believe our data set provides a great resource for additional modelling studies and could help to predict immunity and seroprevalence scenarios during future pandemics.

## Acknowledgements

This effort was supported by the Serological Sciences Network (SeroNet) in part with Federal funds from the National Cancer Institute, National Institutes of Health, under Contract No. 75N91019D00024, Task Order No. 75N91021F00001. The content of this publication does not necessarily reflect the views or policies of the Department of Health and Human Services, nor does mention of trade names, commercial products or organizations imply endorsement by the U.S. Government. This work was also partially funded by the Centers of Excellence for Influenza Research and Surveillance (CEIRS, contract # HHSN272201400008C), the Centers of Excellence for Influenza Research and Response (CEIRR, contract # 75N93021C00014), by the Collaborative Influenza Vaccine Innovation Centers (CIVICs contract # 75N93019C00051) and by institutional funds.

## Author contributions

F.K., V.S., H.v.B., A.G. and E.M.S. conceived and designed the study. H.v.B. wrote and maintained the Institutional Review Board protocol. J.T., A.R., D.B., G.S., S.M., M.F., T.Y., and L.S. performed the serological assays. J.M.C. and F.K. supervised serological assays. D.B., S.M, and B.M. collected, organized and aliquoted plasma samples. G.S. produced and purified recombinant proteins. J.M.C., J.T., B.M., and A.L.W. collected and analyzed the data. J.M.C., B.M., and D.F. generated figures. J.M.C. and A.L.W. wrote original manuscript draft. All authors edited and approved the manuscript.

---

**Supplementary Figure 1.**
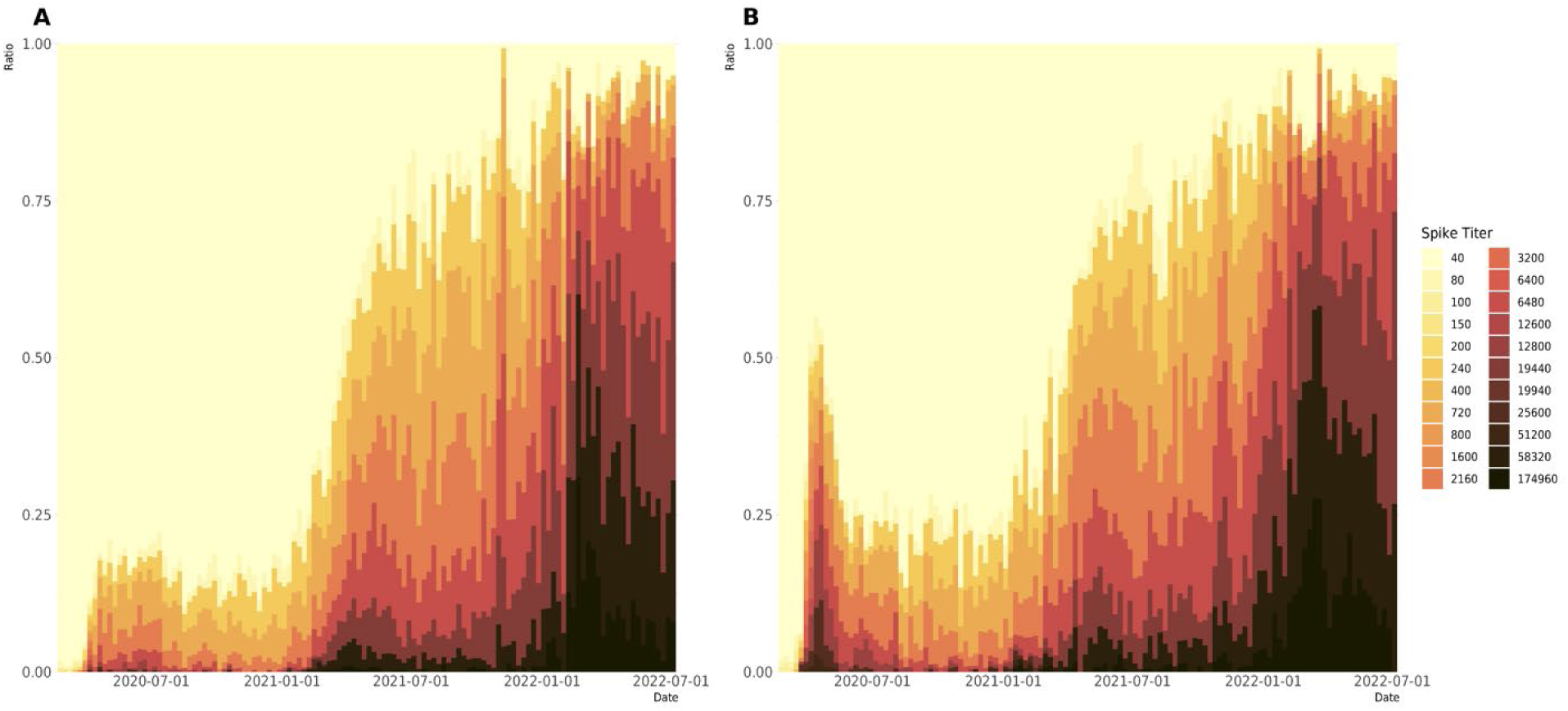
Distribution of SARS-CoV-2 spike antibody titers during the serosurvey period. SARS-CoV-2 spike antibody titers in the Routine Care (left column), and Urgent care (right column) groups measured between February 9^th^ 2020 to July 18^th^ 2022. Weekly counts are presented per single column. Color gradient depicts the range of antibody titers.

**Supplementary figure 2.**
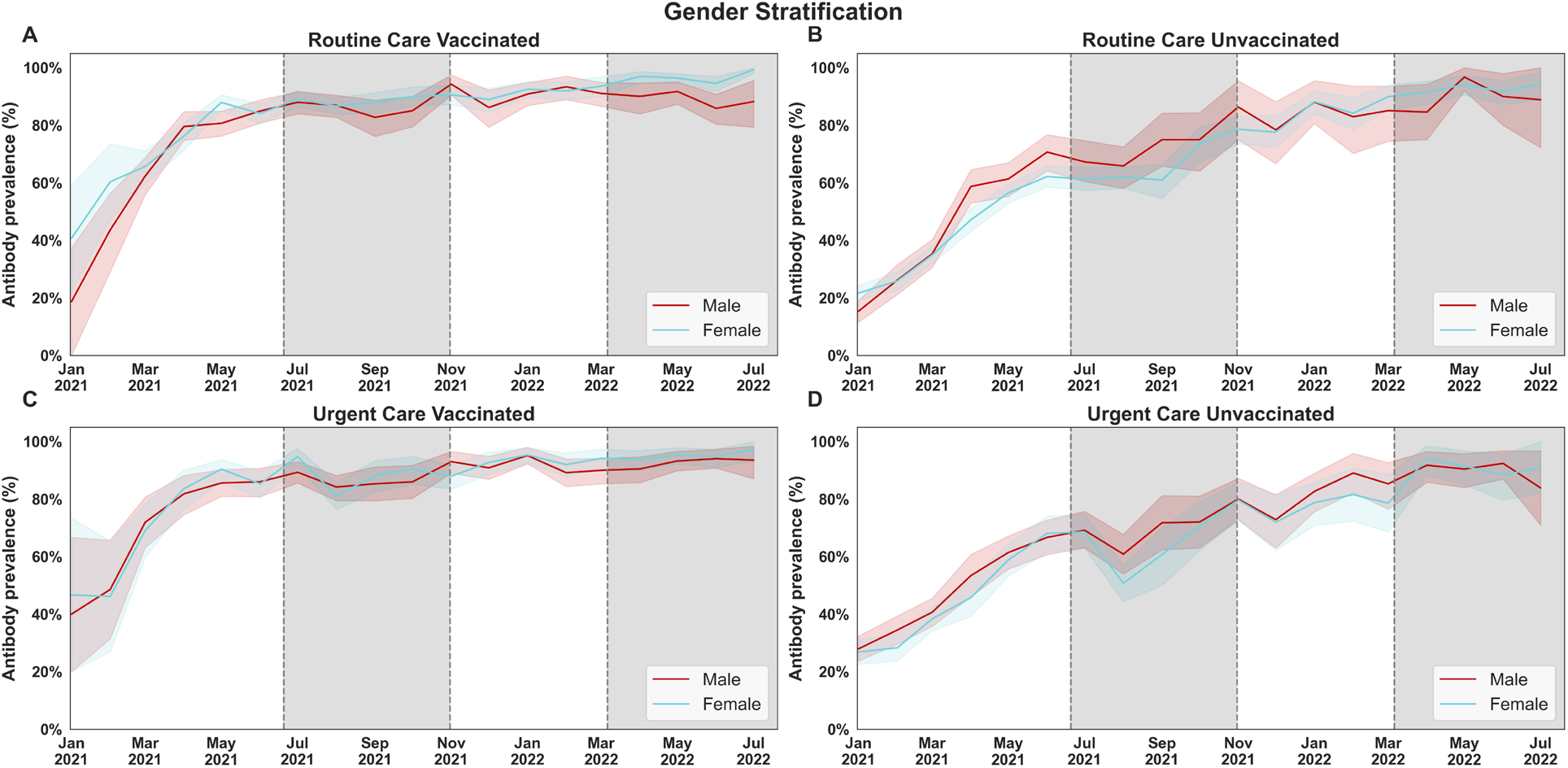
SARS-CoV-2 spike antibody prevalence stratified by sex and vaccination status. Antibody prevalence in the Routine Care (top), and Urgent Care (bottom) groups, measured between February 9^th^ 2020 to July 18^th^ 2022, in vaccinated (left) and unvaccinated (right) inividuals. Charts are fitted with a bootstrapped confidence interval based on random resampling of results to compute an estimate of the 95% confidence interval of the mean. Vaccination status was assessed at time of sample collection and does not reflect vaccination rates in NYC or within our patient population.

**Supplementary figure 3.**
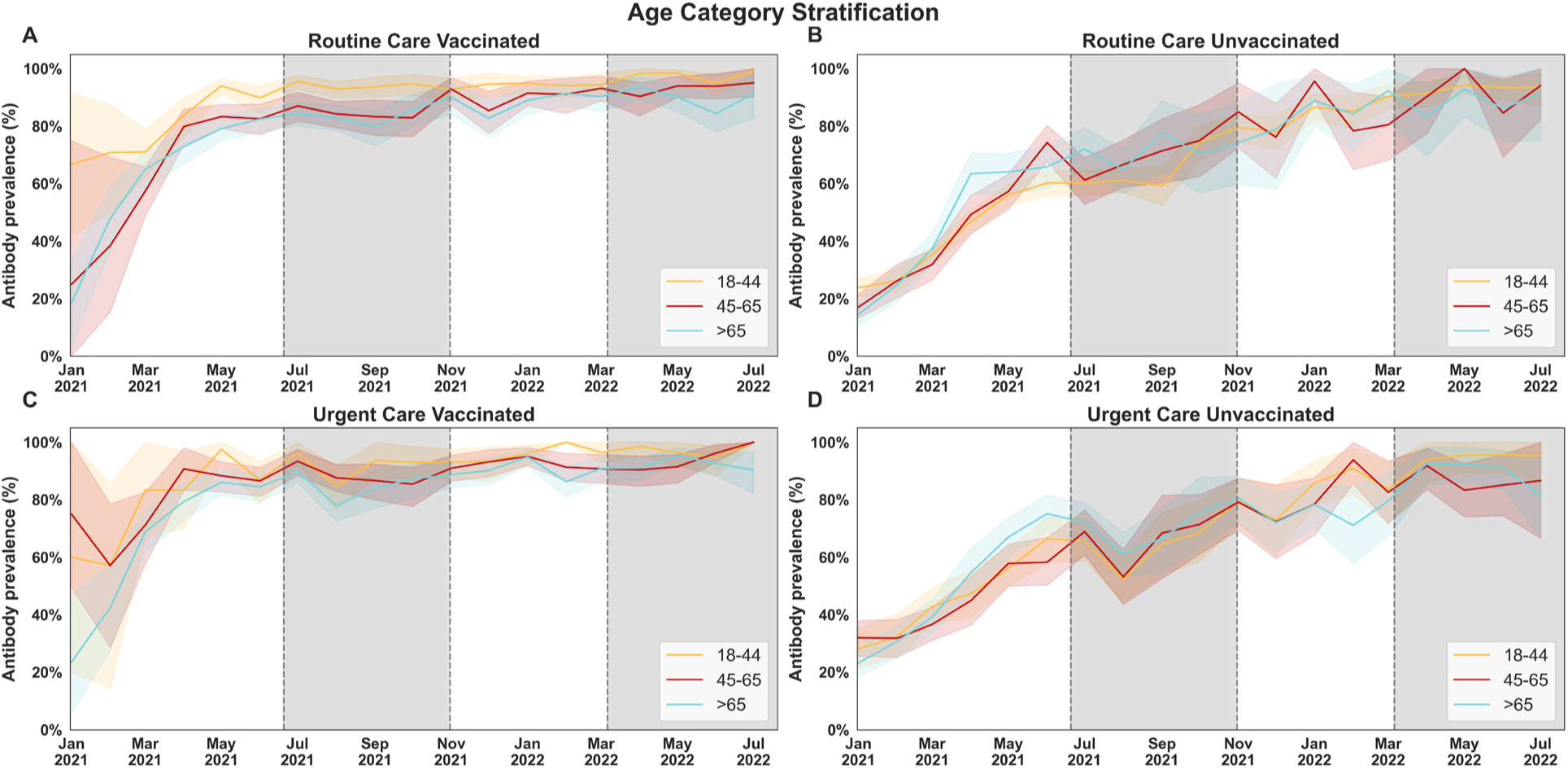
SARS-CoV-2 spike antibody prevalence stratified by age and vaccination status. Antibody prevalence in the Routine Care (top), and Urgent Care (bottom) groups, measured between February 9^th^ 2020 to July 18^th^ 2022 in individuals with differen categorical age levels (18-44, 45-65, and >65 yeal old) in vaccinated (left) and unvaccinated (right) inividuals. Charts are fitted with a bootstrapped confidence interval based on random resampling of results to compute an estimate of the 95% confidence interval of the mean. Vaccination status was assessed at time of sample collection and does not reflect vaccination rates in NYC or within our patient population.

**Supplementary figure 4.**
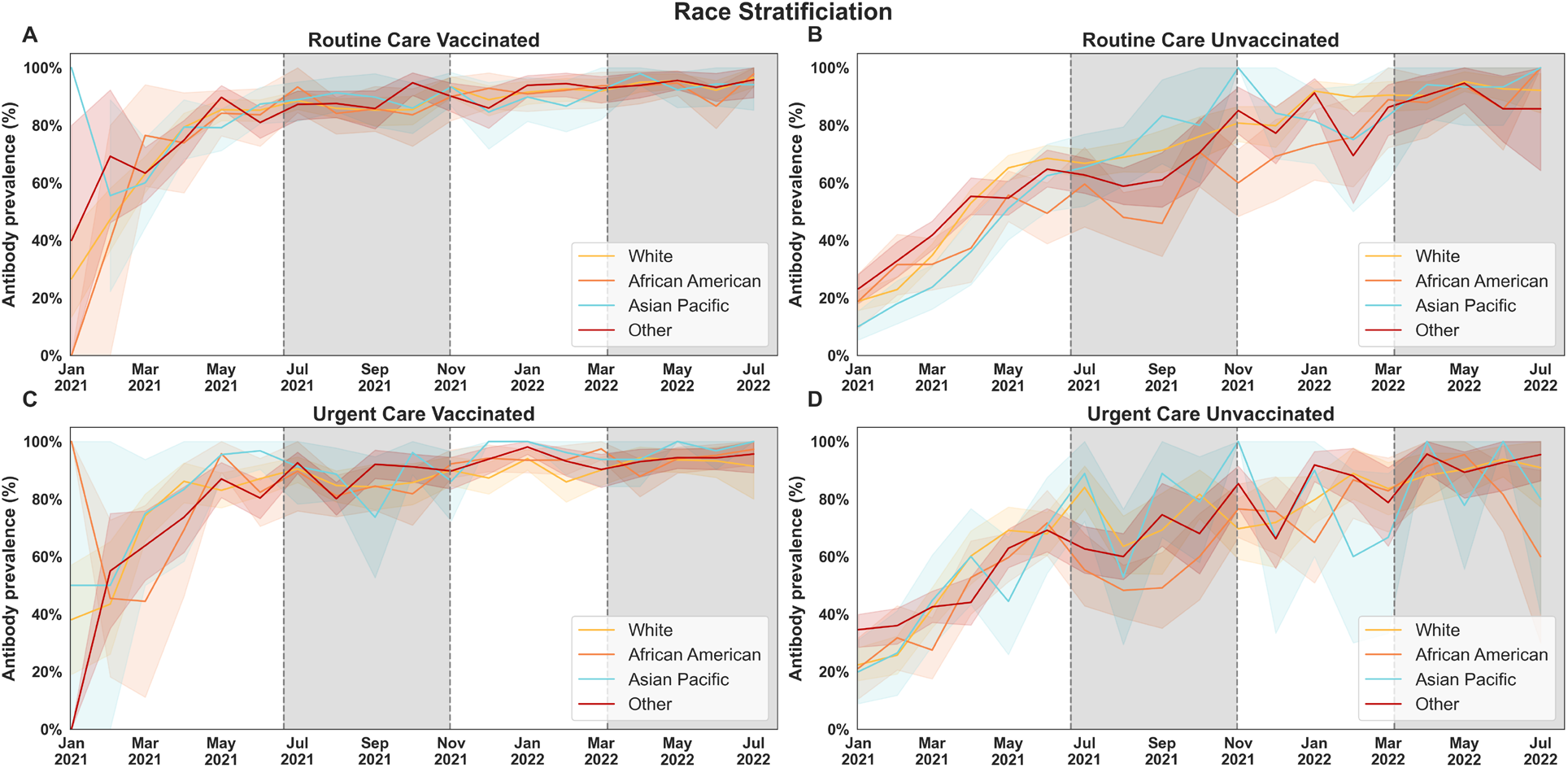
SARS-CoV-2 spike antibody prevalence stratified by race and vaccination status. Antibody prevalence in the Routine Care (top), and Urgent Care (bottom) groups, measured between February 9^th^ 2020 to July 18^th^ 2022 in individuals of different racial categories (White, African American, Asian and Pacific islander, Other, unknown) in vaccinated (left) and unvaccinated (right) individuals. Charts are fitted with a bootstrapped confidence interval based on random resampling of results to compute an estimate of the 95% confidence interval of the mean. Vaccination status was assessed at time of sample collection and does not reflect vaccination rates in NYC or within our patient population.

**Supplementary figure 5.**
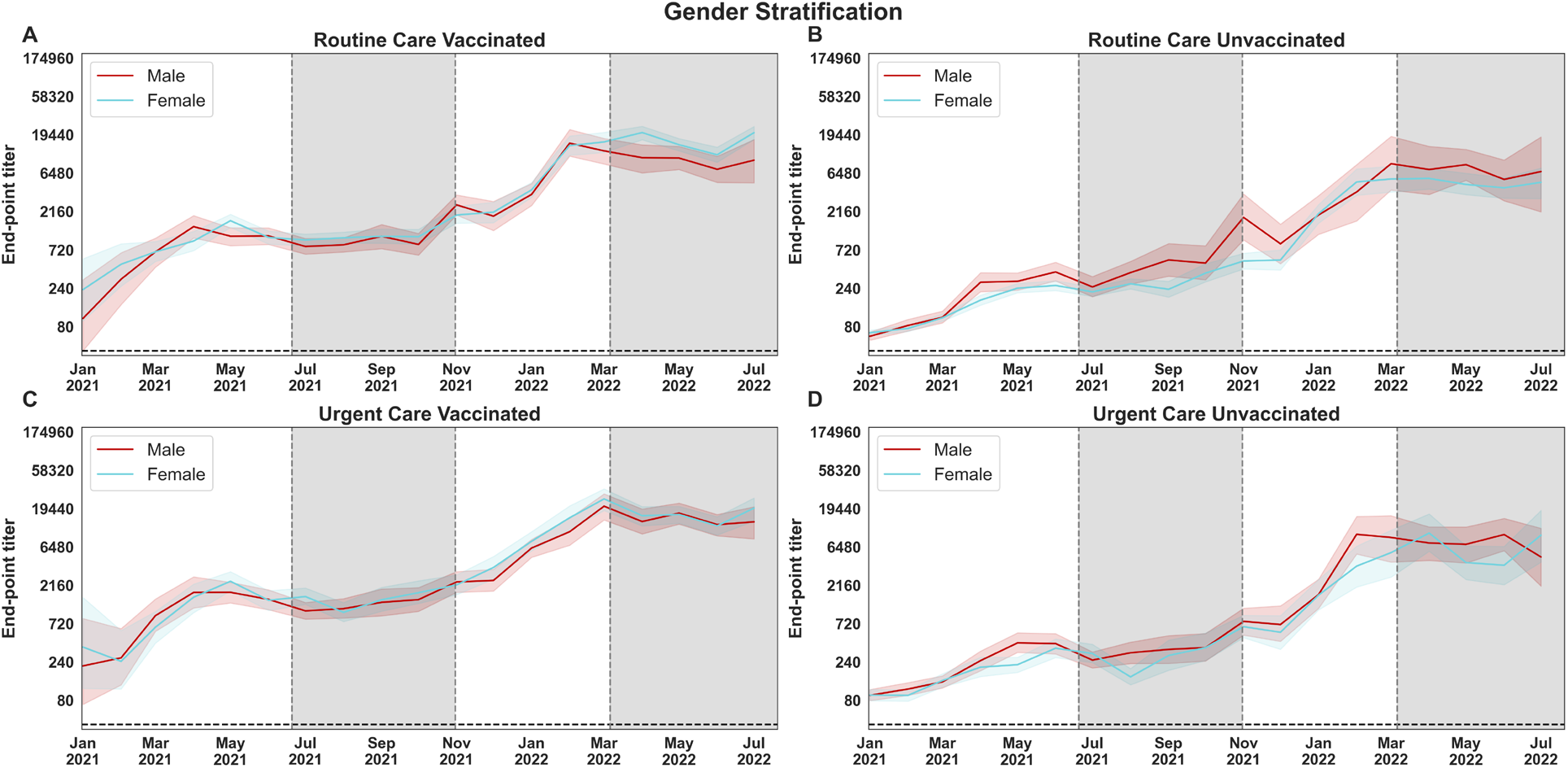
SARS-CoV-2 anti-spike antibody titers stratified by sex and vaccination status. Antibody prevalence in the Routine Care (top), and Urgent Care (bottom) groups, measured between February 9^th^ 2020 to July 18^th^ 2022, in vaccinated (left) and unvaccinated (right) inividuals. Charts are fitted with a bootstrapped confidence interval based on random resampling of results to compute an estimate of the 95% confidence interval of the mean. Vaccination status was assessed at time of sample collection and does not reflect vaccination rates in NYC or within our patient population.

**Supplementary figure 6.**
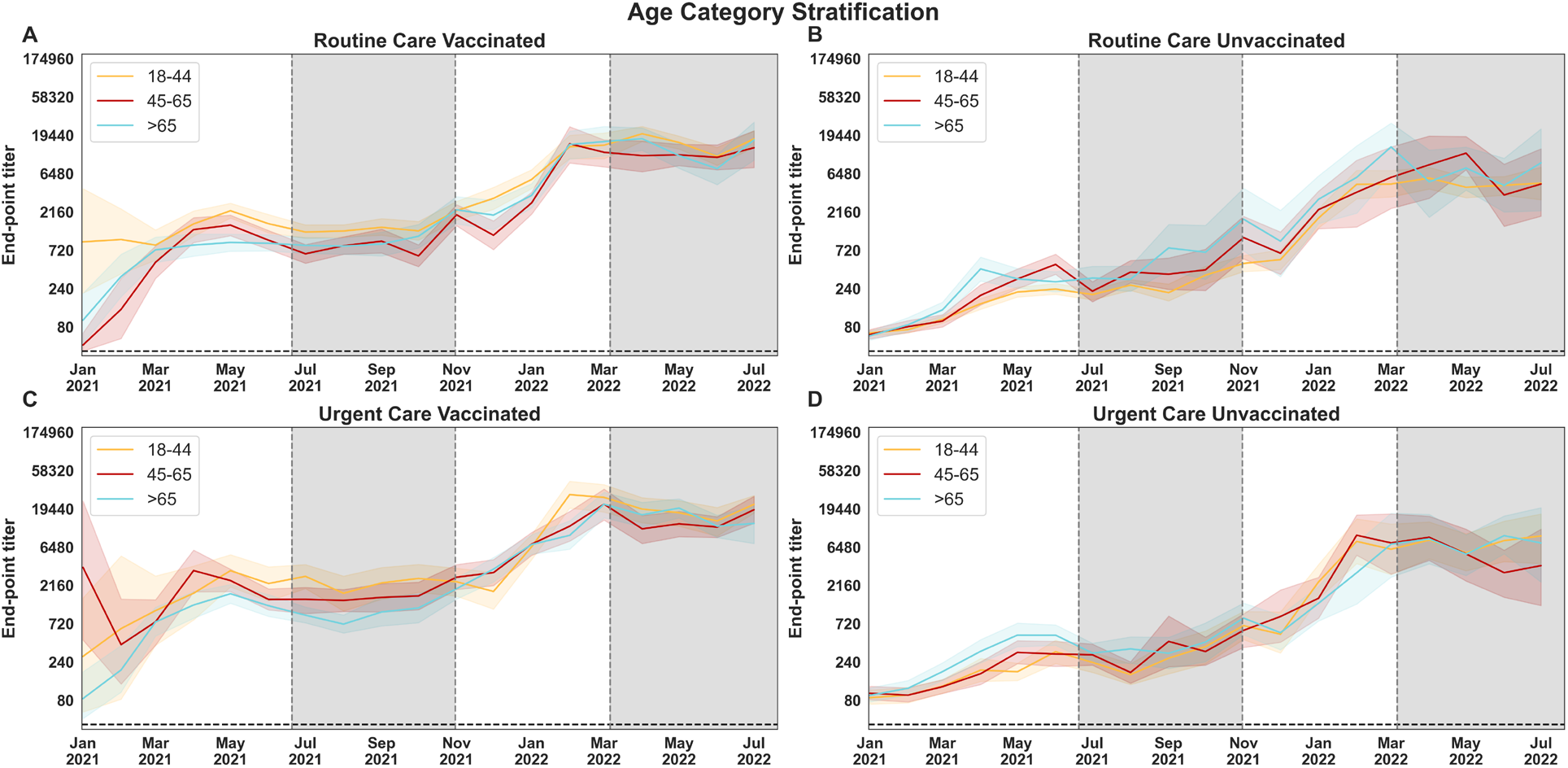
SARS-CoV-2 anti-spike antibody titers stratified by age and vaccination status. Antibody prevalence in the Routine Care (top), and Urgent Care (bottom) groups, measured between February 9^th^ 2020 to July 18^th^ 2022 in individuals with differen categorical age levels (18-44, 45-65, and >65 yeal old) in vaccinated (left) and unvaccinated (right) inividuals. Charts are fitted with a bootstrapped confidence interval based on random resampling of results to compute an estimate of the 95% confidence interval of the mean. Vaccination status was assessed at time of sample collection and does not reflect vaccination rates in NYC or within our patient population.

**Supplementary figure 7.**
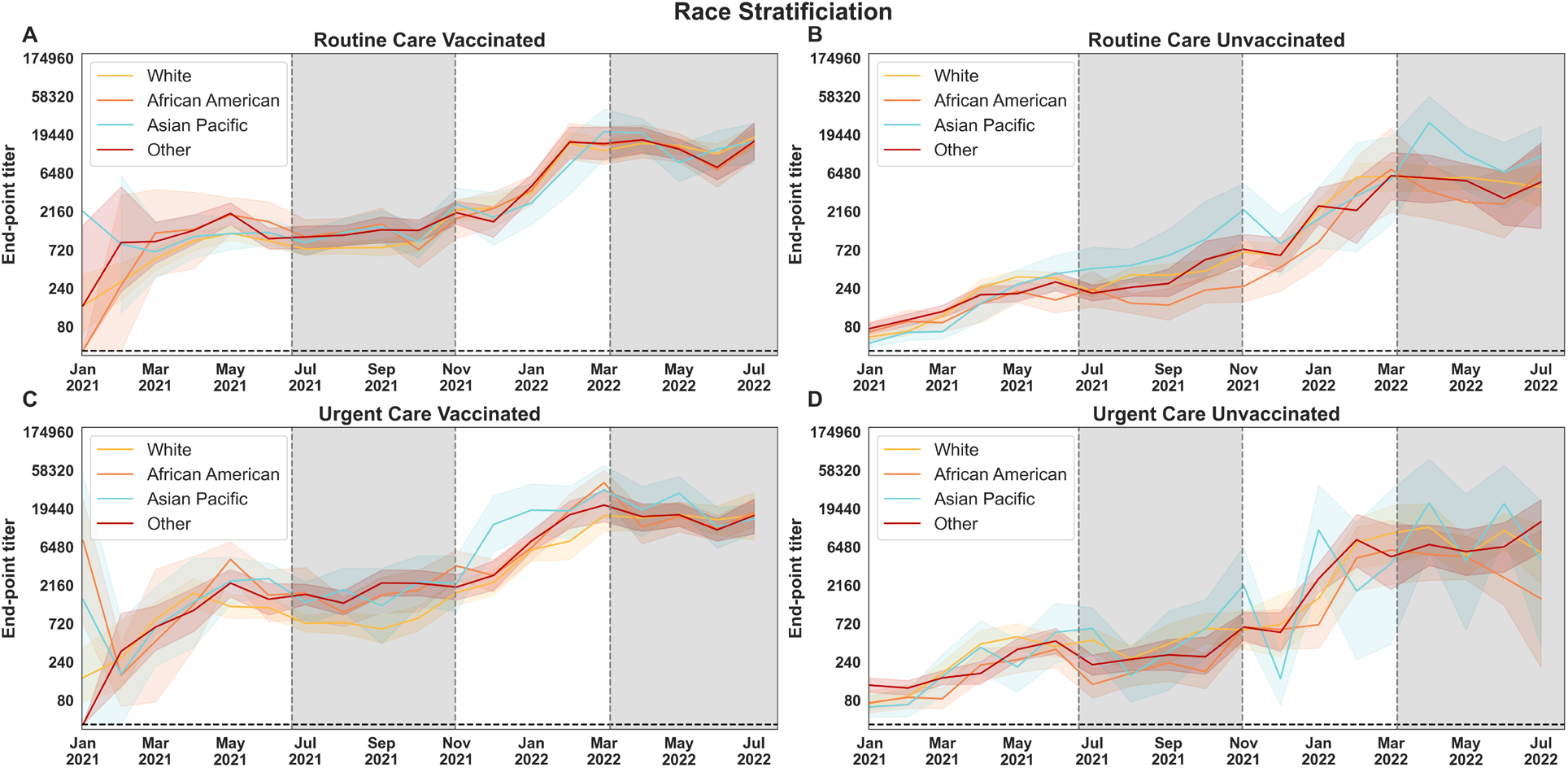
SARS-CoV-2 anti-spike antibody titers stratified by race and vaccination status. Antibody prevalence in the Routine Care (top), and Urgent Care (bottom) groups, measured between February 9^th^ 2020 to July 18^th^ 2022 in individuals of different racial categories (White, African American, Asian and Pacific islander, Other, unknown) in vaccinated (left) and unvaccinated (right) individuals. Charts are fitted with a bootstrapped confidence interval based on random resampling of results to compute an estimate of the 95% confidence interval of the mean. Vaccination status was assessed at time of sample collection and does not reflect vaccination rates in NYC or within our patient population.

## References

1. Thompson, C.N. et al. COVID-19 Outbreak - New York City, February 29-June 1, 2020. MMWR Morb Mortal Wkly Rep 69, 1725-1729 (2020).

2. Dalva-Baird, N.P., Alobuia, W.M., Bendavid, E. & Bhattacharya, J. Racial and ethnic inequities in the early distribution of U.S. COVID-19 testing sites and mortality. Eur J Clin Invest 51, e13669 (2021).

3. Carabelli, A.M. et al. SARS-CoV-2 variant biology: immune escape, transmission and fitness. Nat Rev Microbiol 21, 162–177 (2023).

4. Hou, Y.J. et al. SARS-CoV-2 D614G variant exhibits efficient replication ex vivo and transmission in vivo. Science 370, 1464–1468 (2020).

5. Korber, B. et al. Tracking Changes in SARS-CoV-2 Spike: Evidence that D614G Increases Infectivity of the COVID-19 Virus. Cell 182, 812–827 e819 (2020).

6. Carreno, J.M. et al. Evidence for retained spike-binding and neutralizing activity against emerging SARS-CoV-2 variants in serum of COVID-19 mRNA vaccine recipients. EBioMedicine 73, 103626 (2021).

7. Lucas, C. et al. Impact of circulating SARS-CoV-2 variants on mRNA vaccine-induced immunity. Nature 600, 523–529 (2021).

8. Stadlbauer, D. et al. Repeated cross-sectional sero-monitoring of SARS-CoV-2 in New York City. Nature 590, 146–150 (2021).

9. Khoury, D.S. et al. Neutralizing antibody levels are highly predictive of immune protection from symptomatic SARS-CoV-2 infection. Nat Med 27, 1205–1211 (2021).

10. Gilbert, P.B. et al. Immune correlates analysis of the mRNA-1273 COVID-19 vaccine efficacy clinical trial. Science 375, 43–50 (2022).

11. Carreno, J.M. et al. Longitudinal analysis of severe acute respiratory syndrome coronavirus 2 seroprevalence using multiple serology platforms. iScience 24, 102937 (2021).

12. Singh, G. et al. Binding and avidity signatures of polyclonal sera from individuals with different exposure histories to SARS-CoV-2 infection, vaccination, and Omicron breakthrough infections. J Infect Dis (2023).

13. Schimmel, J., Vargas-Torres, C., Genes, N., Probst, M.A. & Manini, A.F. Changes in alcohol-related hospital visits during COVID-19 in New York City. Addiction 116, 3525–3530 (2021).

14. Zhang, J.J., Dong, X., Liu, G.H. & Gao, Y.D. Risk and Protective Factors for COVID-19 Morbidity, Severity, and Mortality. Clin Rev Allergy Immunol 64, 90–107 (2023).

15. Aziz, H. et al. Antibody Response to SARS-CoV-2 in Relation to the Contributing Factors in COVID-19 Patients. Viral Immunol 35, 142–149 (2022).

16. Masters, N.B. et al. Social distancing in response to the novel coronavirus (COVID-19) in the United States. PLoS One 15, e0239025 (2020).

17. Currie, D. & Wiesenberg, S. Promoting women’s health-seeking behavior: research and the empowerment of women. Health Care Women Int 24, 880–899 (2003).

18. De, P.K. Beyond race: Impacts of non-racial perceived discrimination on health access and outcomes in New York City. PLoS One 15, e0239482 (2020).

19. Spiegelman, D. & Hertzmark, E. Easy SAS calculations for risk or prevalence ratios and differences. Am J Epidemiol 162, 199–200 (2005).

20. Warren, C.M. et al. Assessment of Allergic and Anaphylactic Reactions to mRNA COVID-19 Vaccines With Confirmatory Testing in a US Regional Health System. JAMA Netw Open 4, e2125524 (2021).

21. Department, N.Y.C.H. COVID-19: Data. 2023 [cited 2023 May 25th]Available from: https://www.nyc.gov/site/doh/covid/covid-19-data.page

22. Clarke, K.E.N. et al. Seroprevalence of Infection-Induced SARS-CoV-2 Antibodies - United States, September 2021-February 2022. MMWR Morb Mortal Wkly Rep 71, 606-608 (2022).

23. Do, D.P. & Frank, R. Unequal burdens: assessing the determinants of elevated COVID-19 case and death rates in New York City’s racial/ethnic minority neighbourhoods. J Epidemiol Community Health (2020).

24. Lopez, L., Hart, L.H. & Katz, M.H. Racial and Ethnic Health Disparities Related to COVID-19. JAMA 325, 719–720 (2021).

25. Honein, M.A. et al. Summary of Guidance for Public Health Strategies to Address High Levels of Community Transmission of SARS-CoV-2 and Related Deaths, December 2020. MMWR Morb Mortal Wkly Rep 69, 1860-1867 (2020).

26. Lieberman-Cribbin, W., Galanti, M. & Shaman, J. Socioeconomic Disparities in Severe Acute Respiratory Syndrome Coronavirus 2 Serological Testing and Positivity in New York City. Open Forum Infect Dis 8, ofab534 (2021).

27. A., C.D. Select New York state phase 1b groups are now eligible for COVID-19 vaccination. 2021 [cited 2023 August 25th]Available from: https://www.nyc.gov/assets/doh/downloads/pdf/han/advisory/2021/covid-19-vaccine-1b-eligible-01112021.pdf

28. Carreno, J.M., Singh, G., Simon, V., Krammer, F. & group, P.V.I.s. Bivalent COVID-19 booster vaccines and the absence of BA.5-specific antibodies. Lancet Microbe (2023).

29. Carreno, J.M. et al. Activity of convalescent and vaccine serum against SARS-CoV-2 Omicron. Nature 602, 682–688 (2022).

30. Zhang, A. et al. Beyond neutralization: Fc-dependent antibody effector functions in SARS-CoV-2 infection. Nat Rev Immunol, 1–16 (2022).

31. Tarke, A. et al. SARS-CoV-2 vaccination induces immunological T cell memory able to cross-recognize variants from Alpha to Omicron. Cell 185, 847–859.e811 (2022).

